# The implementation of a falls observational tool in a palliative care setting: a mixed methods study

**DOI:** 10.64898/2026.08.13.26360364

**Authors:** Colette Parfitt, Emma Kirk, Sarah Stanley, Amara Callistus Nwosu

**Affiliations:** Marie Curie North West, Liverpool, Merseyside, UK; Marie Curie North West, Liverpool, Merseyside, UK Liverpool John Moores University, Liverpool, UK, Palliative Care Unit, Liverpool University Hospitals NHS Foundation Trust, Liverpool, UK; Lancaster Medical School, Lancaster University, Lancaster, UK. Marie Curie North West, Liverpool, Merseyside, UK, Palliative Care Unit, Liverpool University Hospitals NHS Foundation Trust, Liverpool, UK

**Keywords:** Palliative care, accidental falls, risk assessment, qualitative research, hospices, patient safety

## Abstract

**Background:** Falls are a major safety concern in healthcare. Palliative patients are particularly vulnerable due to complex symptoms and rapid physical decline. Standard falls risk assessment tools designed for acute settings rely on static scores and lack efficacy in hospice environments. The Falls Early Warning Score (FEWS) is an observational tool developed to address the specific needs of palliative patients; however, evidence describing its use and impact remains limited.

**Aims:** To explore staff views regarding the implementation, utility, and benefits of the FEWS tool in a specialist palliative care inpatient unit.

**Methods:** A mixed-methods study was conducted at a UK hospice. Healthcare professionals using FEWS completed an electronic questionnaire on confidence, practice, and barriers. Findings informed semi-structured, face-to-face interviews. Qualitative data were evaluated using reflexive thematic analysis.

**Results:** Eleven staff completed the questionnaire and five joined interviews. Three major themes emerged: (1) Education: Staff preferred 1:1 training and dedicated user guides; (2) Location and format: Electronic formats aided data collection, while paper charts offered better bedside accessibility, and (3) Recognised benefits: FEWS prompted safe staffing, highlighted variable patient presentation, and mitigated the emotional and physical impact of falls.

**Conclusions:** Integrating bespoke falls risk assessment tools into palliative care is feasible and highly acceptable. Customised tools like FEWS empower staff and support dynamic clinical decision-making. Further research is needed to evaluate their clinical efficacy in reducing falls.

**KEY MESSAGES:** *What is already known on this topic:*

- Falls are one of the most frequently recorded safety incidents in healthcare, and palliative care patients are at a high risk of falling due to fluctuating physical decline and complex symptom burdens.
- Standard falls observational assessment tools are primarily designed for acute hospital settings and lack efficacy in hospice environments because they rely on fixed, static risk scores.

*What this study adds:*

- Bespoke falls observational assessment tools (such as the FEWS chart) are highly acceptable to hospice staff and are perceived to improve falls management by prompting safe staffing levels and highlighting rapid patient deterioration.
- The successful implementation of new clinical tools in palliative care requires continuous, tailored educational support (e.g., 1:1 training and dedicated user guides) and careful consideration of bedside accessibility.

*How this study might affect research, practice or policy:*

- The findings support the evidence base for developing and integrating customised falls management solutions into specialist palliative care clinical practice.
- This study highlights the need for future prospective research to determine the quantitative clinical effectiveness of the FEWS tool in reducing the actual incidence of falls.

## BACKGROUND

Falls in healthcare settings are a major safety concern because they cause severe physical injuries (such as fractures and head trauma), trigger psychological fear that reduces mobility, and lead to prolonged hospital stays or even death.[1, 2] In UK hospitals, falls are the most recorded safety incident, with over 240,000 inpatient falls recorded annually in England and Wales. Consequently, falls management and prevention is a top priority across all healthcare settings.[3] Falls are particularly common in people with palliative care needs due to factors such as declining physical health, impaired mobility and balance, nutritional problems, and symptoms such as pain.[4, 5, 6, 7] Studies report a falls incidence of approximately 10% in palliative care units, with a falls rate of 5.8 to 16.9 per 1000 patient bed days.[8] Furthermore, NICE guidelines suggest that all palliative care patients in an inpatient care setting are at risk of falls, and it is estimated that falls cost the NHS more than £2.3 billion per year.[9]

To improve care and reduce the risk of patients falling, healthcare professionals typically use falls observational assessment tools,[10] such as the ‘Avoiding Falls Level of Observation Assessment Tool’ (AFLOAT).[11] However, while tools like AFLOAT were designed to support the assessment of patients admitted to acute hospital trusts, evidence suggests they lack efficacy for people receiving care in specialist palliative care units.[4] A proposed reason for why standard falls assessment charts are less efficacious in palliative care is their reliance on fixed, static risk-score assessments, which are primarily designed for acute rehabilitation.[12] In contrast, palliative care requires a highly individualised, dynamic approach that balances patient autonomy and quality-of-life against rapidly fluctuating and (usually) irreversible physical deterioration.[13]

To overcome the challenges associated with non-palliative care charts, we developed the Falls Early Warning Score (FEWS) chart, an observational clinical assessment tool designed specifically for hospice and palliative care settings (Supplementary file – FEWS Early Warning Score Form).[14] The FEWS chart uses a simple scoring system paired with clinical action prompts. This helps nursing staff evaluate a palliative patient’s fall risk and determine the appropriate level of supervision.[15] Unlike acute hospital trusts, which often rely on standard risk assessment tools that may not be suitable for end-of-life care, the FEWS chart focuses on the specific complex needs of hospice patients. Although the FEWS chart is used in practice throughout palliative care services in England, there is little evidence regarding staff views on using this tool in clinical practice, or its usefulness in supporting clinical decisions to manage falls risk. A better understanding of staff views regarding their confidence and engagement in using FEWS may help to identify the barriers and facilitators to its implementation, evaluate its perceived impact, and determine how it can be best used to reduce falls risk in palliative care inpatients.

## AIMS

To explore and understand staff views about the benefit of using a palliative care falls observational assessment tool to identify people at risk of falling in a specialist palliative care inpatient unit.

## METHODS

### Study design

This study was conducted in two phases. Firstly, we conducted an electronic questionnaire of staff to understand their views on using the FEWs chart as part of a falls management intervention in a specialist inpatient palliative care unit. Secondly, we conducted semi-structured interviews with staff to understand their perspectives on using the FEWS chart in palliative care practice. We adopted a qualitative descriptive and interpretive framework[16] to capture staff experiences and understand the underlying context of their feedback. This approach allowed us to thoroughly explore healthcare professionals’ insights and generate clinically actionable findings.

### Study Setting

The study was conducted in a hospice in the North West of England, a specialised health care facility providing care for individuals in the advanced stages of a terminal illness or approaching the end of their lives. The hospice provides various services, including a 15-bed inpatient unit, day services, outpatient clinics, community outreach, patient and family support, and bereavement services.

### Questionnaire and Interview Guide Development

We designed a questionnaire using themes from our literature review and built it on Microsoft Forms (Supplementary File - FEWS Questionnaire and electronic consent). The questionnaire consisted mainly of multiple-choice and Likert-scale questions, along with optional free-text fields. Hospice-based patient and public representatives reviewed the initial draft, and we refined the questions based on their feedback. Following the completion of the electronic questionnaire, the preliminary outcomes were analysed by the research team to iteratively develop and refine the questions for the semi-structured interviews. Because the questionnaire responses highlighted clinical trends, practical challenges, and variations in confidence, the interview guide was purposefully tailored to explore these specific questionnaire outcomes in greater depth.

The resulting interview guide consisted of open-ended questions designed to elicit detailed narratives about staff experiences, divided into six domains: demographics and routine practice, accessibility and format of the tool, time constraints and implementation challenges, education and support needs, multidisciplinary team (MDT) involvement, and the overall clinical impact. Participants were asked to evaluate the practicalities of paper versus electronic formats and to reflect on the time taken to complete both initial and regular FEWS assessments. Furthermore, we explored participants’ personal experiences with FEWS education and asked them to provide real-world clinical examples of patient falls, reflecting on the emotional and physical impacts and how the implementation of FEWS might positively change these scenarios. A summary of the interview guide domains and example topics is provided in Table 1, and the complete guide is available in the supplementary file (Supplementary file - Interview guide).

**Table 1:** Summary of Semi-Structured Interview Guide Domains.

| Interview Domain | Key Topics and Example Questions Explored |
| --- | --- |
| Demographics & Routine Practice | Participant's clinical role, time in the role, and whether they routinely complete the FEWS chart. |
| Accessibility & Format | The most popular methods for completing FEWS (e.g., paper vs. electronic), including the respective advantages, disadvantages, and suggestions for making access easier. |
| Time & Implementation Challenges | The time required to complete initial and regular FEWS assessments, practical challenges faced on the ward, and the positives of completing the assessments. |
| Education & Support | Personal education received regarding FEWS, preferred future educational formats (e.g., 1:1 sessions, group presentations), and views on implementing a step-by-step user guide. |
| MDT Involvement | How other members of the multidisciplinary team should be involved, the importance of cross-team communication, and incorporating FEWS into MDT meetings. |
| Clinical Impact & Patient Outcomes | Real-world clinical examples of patient falls, the emotional/physical impact on patients and staff, and how FEWS might influence patient care if made an essential part of admission assessments. |

### Sampling and Recruitment

We collected study data between January to April 2025. We sampled clinical staff of varying roles and experience levels from a specialist hospice inpatient unit in North West England. To be eligible, participants had to be adults (aged ≥18) with clinical experience using or implementing falls observational assessment tools (such as the FEWS chart) and capable of providing informed consent and feedback.

We invited all eligible staff to complete the initial electronic questionnaire, which assessed their confidence in using the FEWS tool. All prospective participants received an email containing study information, participant information sheets, and a link to an electronic consent form. The questionnaire consisted of 20 questions and took approximately 10 minutes complete; however, paper-based versions of all documents (including the questionnaire) were made available upon request. During the consent process, staff were also asked to indicate their willingness to also be interviewed. A member of the research team subsequently contacted the participants who expressed an interest to arrange the semi-structured interviews. All interviews were conducted in person by C.P. and E.K. and recorded using Microsoft Teams on a laptop. Both researchers are experienced female occupational therapists with specialist expertise in palliative care and falls management.

### Data Analysis

#### Questionnaire data analysis

Quantitative data obtained from the electronic questionnaire were analysed using Microsoft Forms to generate simple descriptive statistics and frequency distributions. The preliminary outcomes of this questionnaire analysis were subsequently used by the research team to inform and refine the specific topics explored within the semi-structured interviews.

#### Thematic analysis of interview data

The semi-structured interviews were audio-recorded with participants’ consent and transcribed verbatim. Notes were made by the researcher during the interview. Transcripts were not returned to participants. The qualitative data were then evaluated using reflexive thematic analysis. Following the six-phase process proposed by Braun and Clarke,[17] the analysis iteratively progressed through: (1) becoming familiar with the transcribed data, (2) generating initial codes, (3) developing initial themes, (4) reviewing and refining themes, (5) defining and naming the final themes, and (6) writing up the findings.

The research team maintained a reflective journal throughout data collection to interrogate and contextualize the findings during analysis. Coding was systematically conducted by C.P., E.K., and S.S. using both manual techniques (e.g., highlighters, written notes, and hand-drawn thematic maps) and NVivo 12 software. Interviews continued until thematic saturation was reached, at which point no new themes or insights emerged. Participants did not provide feedback on the outcomes of the data analysis. This study has been reported in accordance with the Consolidated Criteria for Reporting Qualitative Research (COREQ) guidelines.[18] The completed COREQ checklist is available as a supplementary file (Supplementary file - COREQ checklist).

## RESULTS

### Questionnaire characteristics

Eleven healthcare professionals participated in the survey. The majority were female and possessed over ten years of service at the hospice (Table 1). Detailed survey responses are provided in Supplementary File 1 and summarized in Table 2.

**Table 2:** Characteristics of the questionnaire participants (n=11).

| Characteristics | Participants, n (%) |
| --- | --- |
| **Profession** |  |
| Registered Nurse | 4 (36%) |
| Healthcare Assistant | 5 (45%) |
| Occupational Therapist | 1 (9%) |
| Therapy assistant | 1 (9%) |

| Number of years working at the hospice |  |
| --- | --- |
| Less than 6 months | 0 |
| 6-12 months | 0 |
| 12 months - 3 years | 0 |
| 3 years -5 years | 3 (27%) |
| 10 years + | 8 (73%) |

Overall, staff felt confident implementing and evaluating the FEWS tool in daily practice. Key reported barriers were time constraints (n=6, 54%), unfamiliarity with accessing the electronic format (n=6, 54%), and mixed confidence in the tool’s preventative value (n=4, 36%). Suggested solutions included increasing staffing (n=6, 54%), encouraging multidisciplinary participation (n=6, 54%), and providing structured training (n=5, 45%).

Participants universally endorsed initiating the FEWS tool following a fall or near miss (n=11, 100%) or when identifying a falls risk (n=11, 100%). Most also recommended its use during clinical deterioration (n=9, 81%) and routine admission (n=8, 72%).

### Qualitative Findings

We interviewed 5 palliative health care professionals, including three (n=3) registered nurses (60%) and two (n=2) healthcare assistants (40%). A total of four (n=4, 80%) were female and one (n=1, 20%) were male. All interviews were conducted face-to-face, in person and lasted approximately 20 minutes (Table 3). Analysis of the semi-structured interviews revealed three major themes regarding staff views on the Falls Early Warning Score (FEWS) tool: (1) education, (2) tool location and format, and (3) recognised benefits (Figure 1).

**Figure 1:**
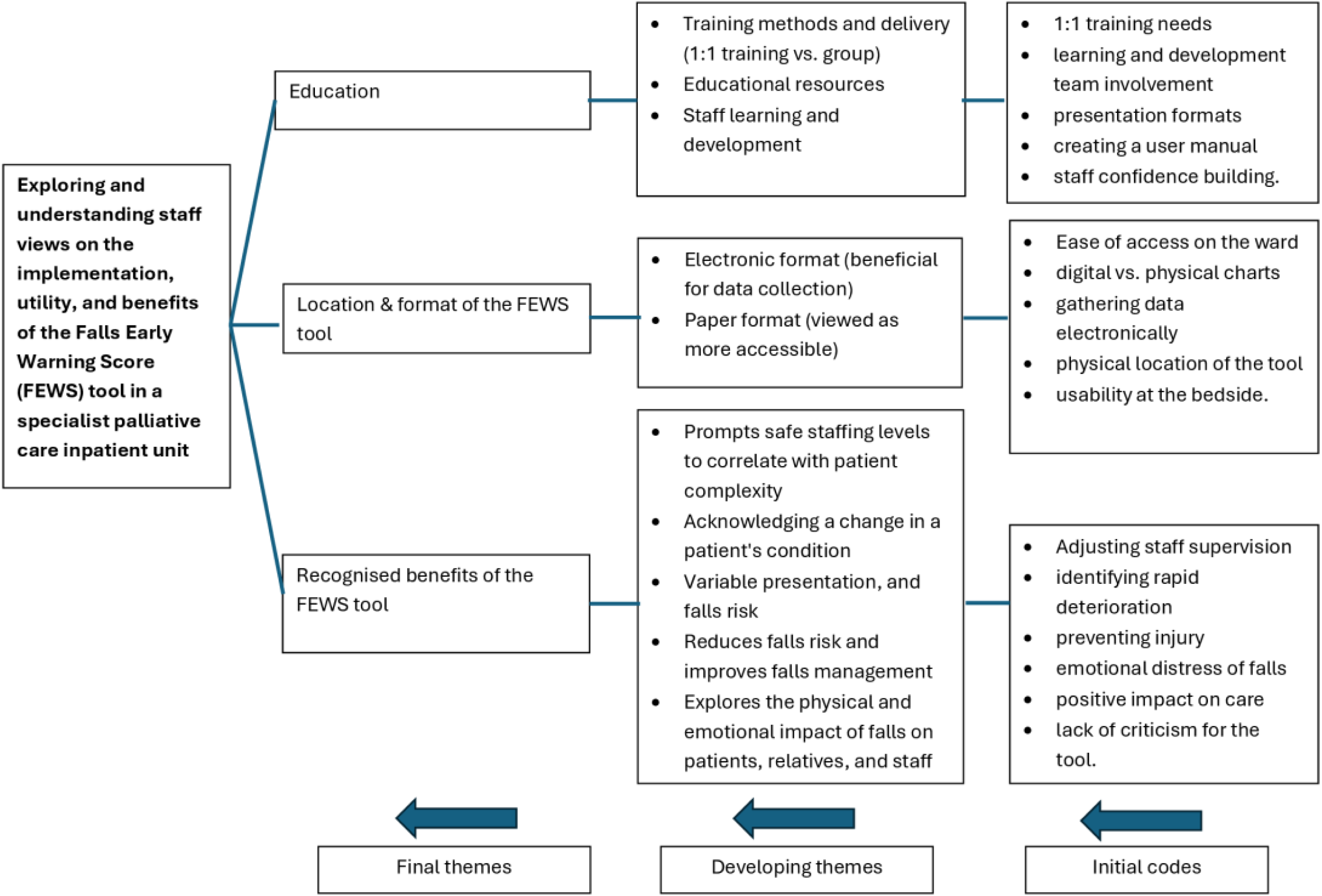
Thematic map showing the 3 main themes.

**Table 3:** Characteristics of the participants interviewed (n=5).

| Characteristics | Participants, n (%) |
| --- | --- |
| **Profession** |  |
| Registered Nurse | 3 (60%) |
| Healthcare Assistant | 2 (40%) |
| **Biological sex** |  |
| Female | 4 (80%) |
| Male | 1 (20%) |
| **Interview method** |  |
| In person | 5 (100%) |
| Online | 0 (0) |

#### Education

Participants highlighted their experiences regarding the training they received to use the FEWS tool, identifying preferences for how this education should be structured and supported. They discussed various methods of training delivery, frequently contrasting the personalised benefits of 1:1 training against the broader reach of group presentations.

*“You know, one to one session is amazing. Yeah. But I also like training when we all together can have a day when we can practice and talk to you girls (therapy team). For 1:1 if we’ve got a new patient and you’re doing assessments first, it would be nice if you can watch how we are doing, which questions you ask here the patients? Yeah, that’d be great. Or go with you. Sometimes if we can to the patient’s house.” (Healthcare Assistant)*

Furthermore, participants emphasised the necessity of ongoing educational support, suggesting that collaboration with the learning and development team and the creation of a dedicated user guide would significantly improve staff confidence and compliance.

*“Because it’s just a prompt, isn’t it? Maybe some guidance on if the score changes because that’s one of the things that doesn’t automatically get reviewed. People sometimes just follow on. They think someone’s an A or a B … things can change… I don’t think that’s always updated as quickly as it could be and is that because they’re not educated to use the tool properly.” (Registered Nurse)*

#### Location and format of the FEWS tool

Participants shared contrasting views regarding how the tool is accessed and used on the ward. Specifically, they weighed the benefits and drawbacks of an electronic format versus a paper format. Some participants acknowledged that an electronic format is beneficial for data collection and overall ward monitoring.

*“Electronic can be completed on the electronic patient records as part of the daily routine… It should be there. But it depends on the person. For me it’s not a problem, but some nurses may find this difficult.” (Healthcare Assistant)*

Others argued that a paper format is more practically accessible and user-friendly for staff working directly at the patient’s bedside.

*“Paper, yeah… I think it’s easy just to pick up and do it at the bedside or the nurse’s station*.

*When it’s electronic, I don’t think we’d be able to log on… because as nurses, we’re not always logged on are we? We’re more hands on.” (Registered Nurse)*

#### Recognised benefits of the FEWS tool

Participants acknowledged several clinical and psychological benefits resulting from use of the FEWS tool. Participants described how FEWS acted as a prompt to ensure safe staffing levels that correlate directly with patient complexity. Furthermore, staff noted that the tool helped staff to acknowledge changes in a patient’s condition, which helped to reduce risk of falls and improve management.

*“People’s condition can change and can be variable can’t it, so from one day to the next, or morning into the afternoon. So, we just highlight that things can change and how you can put something extra in place that you might need later on in the day or the following day… So, from a staffing point of view, it’s a good tool.” (Registered Nurse)*

Participants also described the physical and emotional impact that falls have on patients, their relatives, and the staff themselves, noting that the FEWS tool could potentially help to reduce the distress associated with falls.

*“Oh, that (FEWs) is absolutely important because it prevents patients from accidents because if they have an accident, they need to go to hospital, they feel worse. It is very, very important.” (Healthcare Assistant)*

## DISCUSSION

### Summary of Main Findings

This study highlights the potential of the FEWS tool to enhance falls management in a hospice inpatient setting. Concerning the implementation and use of the FEWS chart in palliative care, our findings demonstrate that it is important to consider the educational support provided to staff, the practical location and format of the tool, and the recognised clinical benefits of using the FEWS tool in practice.

### Importance and Strengths of the Study

Our paper provides useful insight into staff views on fall management in palliative care. Understanding these perspectives is important to ensure that fall assessment tools designed for people with palliative care needs (like FEWS) are implemented successfully and effectively. Furthermore, by addressing staff needs and engagement, our findings enhance the transferability of the tool, to facilitate translation of the learning from this study to other palliative care settings to improve patient safety.

### Comparison to Previous Work

Our study highlights that staff education and development programmes (for example, 1:1 training, collaboration with learning and development teams, and the creation of dedicated user guides) are important for the successful implementation of falls management strategies. Our findings are consistent with previous work which stresses that falls management in palliative care requires a highly individualised, dynamic approach that prioritises patient autonomy whilst acknowledging their condition can rapidly deteriorate.[4] Existing tools (e.g. AFLOAT), are primarily designed for acute clinical environments and rely on fixed, static risk-score assessments.[11] While acute tools often seek a simple prediction of risk, our findings demonstrate that hospice staff require continuous learning and tailored educational support to confidently apply clinical judgement to complex, changing patient needs.

Consistent with previous work, our participants emphasised the importance of the location and format of the assessment tool, specifically describing the benefits of using electronic formats for data collection compared to paper charts.[19, 20] Specifically, electronic data capture offers potential for accurate real-time data capture and analysis, improving assessment, incorporation of patient reported outcome measures, enabling the rapid changes in the health of palliative care patients to be identified and managed.[21, 22] Our findings support previous work which shows that palliative care patients have different needs compared to more stable general hospital patients, as they frequently experience sudden changes in their health that require immediate assessment.[23] Therefore, having an accessible, highly visible tool at the bedside empowers staff to respond in real-time to these rapid deteriorations, as well as to environmental hazards common to hospice layouts (e.g. medical equipment like syringe driver infusion pumps, oxygen cylinders and tubing).

Finally, staff recognised key benefits of the FEWS chart, such as prompting safe staffing levels based on patient complexity and highlighting clinical changes in a timely manner. These features address critical gaps in traditional falls assessment tools, which often fail to account for palliative-specific factors like severe symptom burden (e.g., fatigue and breathlessness).[4] Our findings align with prior research demonstrating that when a tool actively prompts staff to address palliative-specific factors, it improves falls management and reduces physical and emotional distress for patients, families, and healthcare staff.[4, 6, 8]

### Limitations

This study has several limitations. First, the sample size was small and restricted to a single, small hospice, which may limit the generalisability of the findings. The participant demographic was mainly white, and the study was conducted exclusively in the English language; therefore, the views captured may not fully represent diverse cultural backgrounds. Consequently, the data may not be transferable to other healthcare environments, and we are not able to draw conclusions regarding whether the FEWS chart is useful or works in other settings such as patients’ homes, care homes, or acute hospitals. Additionally, because the FEWS chart functions strictly as a risk predictor, this study did not evaluate the physiological or mechanical mechanisms underlying patient falls. Finally, as this specific paper focuses on staff perspectives, it does not provide definitive data on the quantitative or qualitative impacts of the tool in practice.

### Implications for Policy and Practice

Despite these limitations, our work informs current policy and practice by supporting the evidence base for the process of developing and integrating tailored falls management solutions into palliative care clinical practice. We highlight the importance of considering staff needs during the implementation of falls assessment tools, particularly regarding the necessity of having access to educational materials and training to improve clinical management.[24]

### Future Research

Further research is needed to explore how falls management requirements vary for palliative care patients across different care settings. Additionally, studies should examine how fall management procedures and stakeholder needs differ among organisations. Future research should evaluate the quantitative impact of the tool in practice to definitively determine if the FEWS tool works in reducing falls among palliative care patients.

## CONCLUSION

This study shows that integrating customized falls assessment tools (like the FEWS chart) into specialist palliative care is both feasible and acceptable to staff. However, further research is needed to determine whether these tools safely and effectively reduce patient falls across broader clinical settings.

## Author Contributions

### CRediT Author Contributions

- **Colette Parfitt (CP):** Conceptualization, Methodology, Funding Acquisition, Investigation, Project Administration, Formal Analysis, Writing – Original Draft, Writing – Review & Editing.
- **Emma Kirk (EK):** Investigation, Project Administration, Writing – Original Draft, Writing – Review & Editing.
- **Sarah Stanley (SS):** Conceptualization, Methodology, Funding Acquisition, Investigation, Project Administration, Formal Analysis, Supervision, Writing – Original Draft, Writing – Review & Editing.
- **Amara Callistus Nwosu (ACN):** Methodology, Funding Acquisition, Investigation, Project Administration, Supervision, Writing – Original Draft, Writing – Review & Editing.

## Statements and Declarations

### Ethical considerations

Ethical approval for this study was granted by the Faculty of Health and Medicine (FHM) Research Ethics Committee of Lancaster University, Lancaster, UK. FHM REC Reference: FHM-2024-4313-RECR-2.

### Consent to participate

Informed consent to participate was obtained participants prior to their involvement.

### Consent for publication

Participants provided consent for their anonymised data and quotes to appear in this manuscript.

### Data availability

Summaries of the qualitative data generated and analysed during the current study are available in in the manuscript and the supplementary material.

## SUPPLEMENTARY FILES

- FEWS Early Warning Score Form
- FEWS Questionnaire and electronic consent
- Interview guide
- COREQ checklist
- Results of FEWS questionnaire

## Supporting information

Supplementary file - COREQ checklist

Supplementary file - Results of FEWS questionnaire

Supplementary file - FEWS Questionnaire and electronic consent

Supplementary file - Interview guide

Supplementary file - FEWS Early Warning Score Form

