## Supplementary file - COREQ checklist for "The implementation of a falls observational tool in a palliative care setting: a mixed methods study"

### COREQ (COnsolidated criteria for REporting Qualitative research) Checklist

A checklist of items that should be included in reports of qualitative research. You must report the page number in your manuscript where you consider each of the items listed in this checklist. If you have not included this information, either revise your manuscript accordingly before submitting or note N/A.

| Topic | Item No. | Guide Questions/Description | Reported on Page No. |
|---|---|---|---|
| **Domain 1: Research team and reflexivity** | | | |
| *Personal characteristics* | | | |
| Interviewer/facilitator | 1 | Which author/s conducted the interview or focus group? | 10 |
| Credentials | 2 | What were the researcher's credentials? E.g. PhD, MD | 1-2 |
| Occupation | 3 | What was their occupation at the time of the study? | |
| Gender | 4 | Was the researcher male or female? | 10 |
| Experience and training | 5 | What experience or training did the researcher have? | 10 |
| *Relationship with participants* | | | |
| Relationship established | 6 | Was a relationship established prior to study commencement? | 6-7 |
| Participant knowledge of the interviewer | 7 | What did the participants know about the researcher? e.g. personal goals, reasons for doing the research | 6-7 |
| Interviewer characteristics | 8 | What characteristics were reported about the inter viewer/facilitator? e.g. Bias, assumptions, reasons and interests in the research topic | 6-7, 10, 18 |
| **Domain 2: Study design** | | | |
| *Theoretical framework* | | | |
| Methodological orientation and Theory | 9 | What methodological orientation was stated to underpin the study? e.g. grounded theory, discourse analysis, ethnography, phenomenology, content analysis | 7 |
| *Participant selection* | | | |
| Sampling | 10 | How were participants selected? e.g. purposive, convenience, consecutive, snowball | 10 |
| Method of approach | 11 | How were participants approached? e.g. face-to-face, telephone, mail, email | 10 |
| Sample size | 12 | How many participants were in the study? | 11-13 |
| Non-participation | 13 | How many people refused to participate or dropped out? Reasons? | NA |
| *Setting* | | | |
| Setting of data collection | 14 | Where was the data collected? e.g. home, clinic, workplace | 10 |
| Presence of non-participants | 15 | Was anyone else present besides the participants and researchers? | NA |
| Description of sample | 16 | What are the important characteristics of the sample? e.g. demographic data, date | 11-13 |
| *Data collection* | | | |
| Interview guide | 17 | Were questions, prompts, guides provided by the authors? Was it pilot tested? | 9 |
| Repeat interviews | 18 | Were repeat inter views carried out? If yes, how many? | NA |
| Audio/visual recording | 19 | Did the research use audio or visual recording to collect the data? | 10 |
| Field notes | 20 | Were field notes made during and/or after the inter view or focus group? | 11 |
| Duration | 21 | What was the duration of the inter views or focus group? | 13 |
| Data saturation | 22 | Was data saturation discussed? | 11 |
| Transcripts returned | 23 | Were transcripts returned to participants for comment and/or | 11 |

| Topic | Item No. | Guide Questions/Description | Reported on Page No. |
|---|---|---|---|
| | | correction? | |
| **Domain 3: analysis and findings** | | | |
| *Data analysis* | | | |
| Number of data coders | 24 | How many data coders coded the data? | 11 |
| Description of the coding tree | 25 | Did authors provide a description of the coding tree? | 14 |
| Derivation of themes | 26 | Were themes identified in advance or derived from the data? | 13-16 |
| Software | 27 | What software, if applicable, was used to manage the data? | 11 |
| Participant checking | 28 | Did participants provide feedback on the findings? | 11 |
| *Reporting* | | | |
| Quotations presented | 29 | Were participant quotations presented to illustrate the themes/findings? Was each quotation identified? e.g. participant number | 14-16 |
| Data and findings consistent | 30 | Was there consistency between the data presented and the findings? | 11-19 |
| Clarity of major themes | 31 | Were major themes clearly presented in the findings? | 14 |
| Clarity of minor themes | 32 | Is there a description of diverse cases or discussion of minor themes? | 14 |

Developed from: Tong A, Sainsbury P, Craig J. Consolidated criteria for reporting qualitative research (COREQ): a 32-item checklist for interviews and focus groups. *International Journal for Quality in Health Care*. 2007. Volume 19, Number 6: pp. 349 – 357

**Once you have completed this checklist, please save a copy and upload it as part of your submission. DO NOT include this checklist as part of the main manuscript document. It must be uploaded as a separate file.**
