## Supplementary file - Results of FEWS questionnaire for "The implementation of a falls observational tool in a palliative care setting: a mixed methods study"

1. I confirm that I have read the information sheet dated 11.07.24 for the above study. I have had the opportunity to consider the in formation, ask questions and have had these answered satisfactorily. More details

- yes 11
- no 0

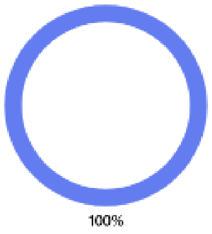


2. I understand what taking part in the study involves More details

- yes 11
- no 0

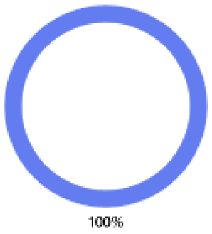


3. I consent voluntarily to be a participant in this study and understand that I can refuse to answer questions. You can withdraw fro m the study at any time, without giving a reason, but we will keep information about you that we already have. More details

- yes 11
- no 0

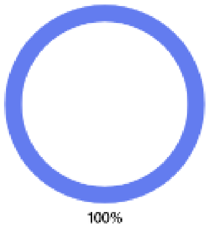


4. I understand who will have access to personal data provided, how the data will be stored and what will happen to the data at the end of the project. More details

- yes 11
- no 0

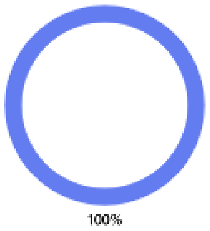

5. I understand that my information may be subject to review by responsible individuals from Lancaster University and/or Marie Curie for monitoring and audit purposes

More details

- yes 11
- no 0

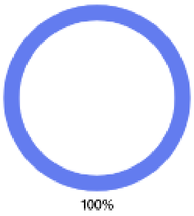


6. I agree for my contact details to be stored for the purpose of contacting me about future studies and I understand that agreeing to be contacted does not oblige me to participate in any further studies

More details

- yes 11
- no 0

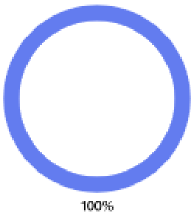


7. I understand that personal data will be retained beyond the duration of the study

More details

- yes 11
- no 0

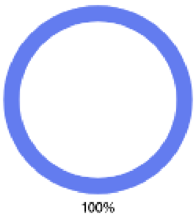


8. I understand that personal data will remain confidential and that all efforts will be made to ensure I cannot be identified in reports or any further outputs

More details

- yes 11
- no 0

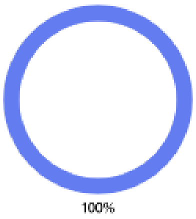


9. I understand that even though all efforts will be made to ensure I cannot be identified, I may be indirectly identifiable when the study findings are disseminated.

More details

- yes 11
- no 0

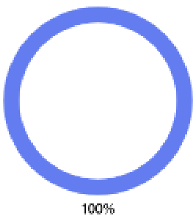

10. I understand the potential risks of being identifiable in reports and any future outputs when the findings of the study are disse minated

More details

| | |
|---|---|
| yes | 11 |
| no | 0 |

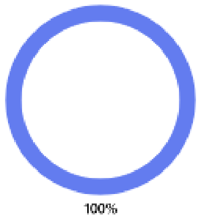


11. I agree to take part in this study

More details

| | |
|---|---|
| yes | 11 |
| no | 0 |

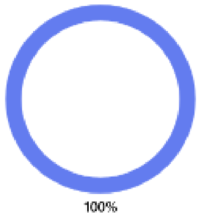


12. What is your role in the inpatient unit?

More details

| | |
|---|---|
| Nursing | 4 |
| Health care assistant | 5 |
| medical | 0 |
| Patient and family support team | 0 |
| Occupational therapy/physiotherapy team | 2 |
| Management | 0 |
| Administration | 0 |
| Other | 0 |
| Other | 0 |

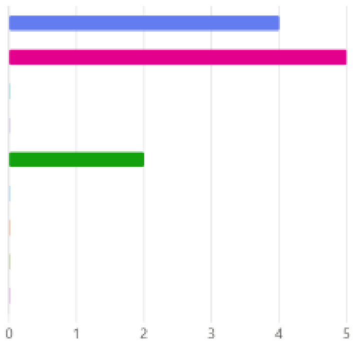


13. How long have you worked on the inpatient unit in Marie Curie Hospice Liverpool

More details

| | |
|---|---|
| Less than 6 months | 0 |
| 6-12 months | 0 |
| 12 months - 3 years | 0 |
| 3 years -5 years | 0 |
| 5 years - 10 years | 3 |
| 10 years + | 8 |

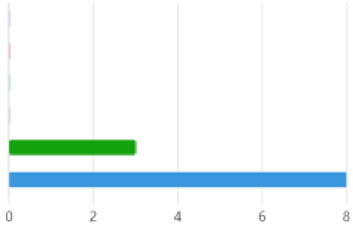

14. On a scale of 1-10 how confident would you feel to implement a FEWS tool as part of inpatient care?

More details

8.18

Average Rating

1 2 3 4 5 6 7 8 9 10

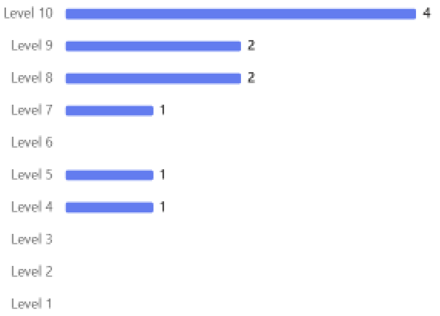


15. How-on a scale of 1-10 how confident would you feel to review a FEWs tool as part of inpatient care?

More details

8.09

Average Rating

1 2 3 4 5 6 7 8 9 10

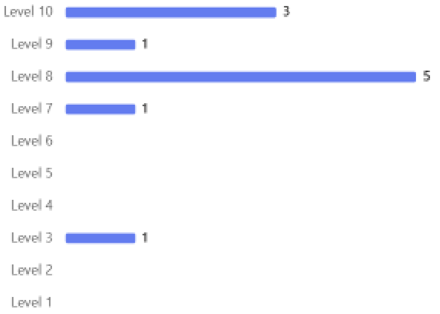


16. What are the barriers to FEWS being used on the ward?
(Select one or more option)

More details

| | |
|---|---|
| Lack of time | 6 |
| Not aware of what the FEWS tool is | 1 |
| Not aware of how to use the FEWS tool | 3 |
| Confidence in the FEWs tool working as a falls prevention tool | 4 |
| Lack of printed forms available | 0 |
| Unware how to access the electronic version of the FEWS tool | 6 |
| Feel the FEWs tool isn't needed | 0 |
| There are no barriers to using the FEWs tool | 1 |
| Other | 3 |

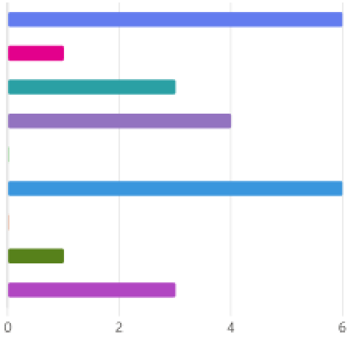


17. If you answered 'other' to question 16, please provide details below

More details

2

Responses

Latest Responses

"n/a"

"n/a"

18. How do you feel that the barriers to implementing FEWs could be overcome?
(Select one or more option)

More details

| | |
|---|---|
| Training on the use of FEWs | 5 |
| Improved staffing levels | 6 |
| Tool more accessible (admin to support printing out forms) | 1 |
| Multidisciplinary involvement in completing the FEWS tool | 6 |
| Written guidance on how to complete | 1 |
| Other | 2 |

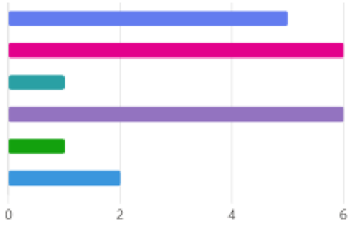


19. If you answered 'other' to question 18, please provide details below

More details

2
Responses

Latest Responses
"explaining properly how it is used"
...

20.
Have you received training on FEWS?

More details

| | |
|---|---|
| Yes | 4 |
| No | 7 |

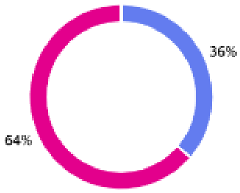


21. How did you access training on FEWs?
(You can select more than one answer)

More details

| | |
|---|---|
| Hospice education sessions | 4 |
| 1:1 training during clinical practice | 0 |
| MDT discussions | 0 |
| Journal clubs | 0 |
| Other | 0 |

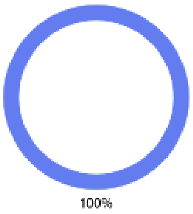


22. If you answered 'other' to the previous question, please provide details below

0 responses submitted

0
Responses

23. Who do you think should be responsible for initiating the FEWs tool? completing and reviewing the tool?
(You can select more than one answer)

More details

| | |
|---|---|
| Occupational therapist | 9 |
| Physiotherapist | 9 |
| Nursing staff | 8 |
| Health care assistants | 8 |
| Medical staff | 8 |
| Other | 2 |

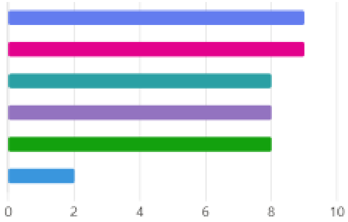

24. If you answered 'other' to the previous question, please provide details below

More details

2
Responses

Latest Responses
...

25. Who do you think should be responsible for completing the FEWs tool?
(You can select more than one answer)

More details

| | |
|---|---|
| Occupational therapist | 11 |
| Physiotherapist | 11 |
| Nursing staff | 10 |
| Health care assistants | 10 |
| Medical staff | 9 |
| Other | 2 |

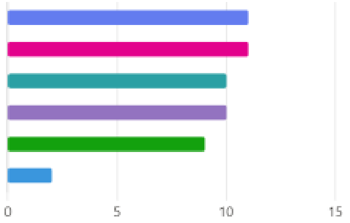


26. If you answered 'other' to the previous question, please provide details below

More details

3
Responses

Latest Responses
"anybody"
...

27. Who do you think should be responsible for reviewing the level of observation required using the FEWs tool?
(You can select more than one answer)

More details

| | |
|---|---|
| Occupational therapist | 11 |
| Physiotherapist | 11 |
| Nursing staff | 10 |
| Health care assistants | 9 |
| Medical staff | 10 |
| Other | 2 |

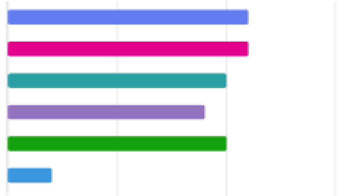

28. If you answered 'other' to the previous question, please provide details below

More details

3
Responses

Latest Responses
"any ahp that looks after patient"
...

29. Do you know when FEWs should be initiated?

More details

| | |
|---|---|
| On admission | 8 |
| Following a fall or near miss | 11 |
| If medical condition detoriates | 9 |
| When risk of fall is identified | 11 |
| Unsure | 0 |

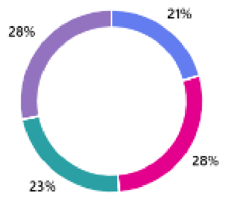


30. Would you be happy for us to contact you to ask you to participate in an interview following this questionnaire?

More details

| | |
|---|---|
| Yes | 7 |
| No | 4 |

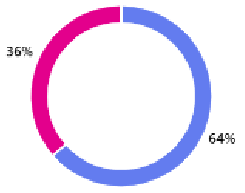
