## Supplementary file - FEWS Questionnaire and electronic consent for "The implementation of a falls observational tool in a palliative care setting: a mixed methods study"

Falls Early Warning Score (FEWS CHART);
The implementation of a falls observational tool and its clinical effectiveness of reducing falls in a palliative care setting.

We have developed this research project with the aim to improve falls management by supporting healthcare professionals to become more confident when making clinical decisions and implementing falls management plans. It is important that we hear your views about using the FEWs chart in clinical practice.

* Required

### Consent to participate

Please read the participant information sheet before providing consent to participate: *INSERT LINK*

1. I confirm that I have read the information sheet dated **INSERT** for the above study. I have had the opportunity to consider the information, ask questions and have had these answered satisfactorily. *

◯ yes

◯ no

2. I understand what taking part in the study involves *

◯ yes

◯ no

3. I consent voluntarily to be a participant in this study and understand that I can refuse to answer questions. You can withdraw from the study at any time, without giving a reason, but we will keep information about you that we already have. *

◯ yes

◯ no

4. I understand who will have access to personal data provided, how the data will be stored and what will happen to the data at the end of the project. *

◯ yes

◯ no

5. I understand that my information may be subject to review by responsible individuals from Lancaster University and/or Marie Curie for monitoring and audit purposes *

◯ yes

◯ no

6. I agree for my contact details to be stored for the purpose of contacting me about future studies and I understand that agreeing to be contacted does not oblige me to participate in any further studies *

◯ yes

◯ no

7. I understand that personal data will be retained beyond the duration of the study *

◯ yes

◯ no

8. I understand that personal data will remain confidential and that all efforts will be made to ensure I cannot be identified in reports or any further outputs *

◯ yes

◯ no

9. I understand that even though all efforts will be made to ensure I cannot be identified, I may be indirectly identifiable when the study findings are disseminated. *

◯ yes

◯ no

10. I understand the potential risks of being identifiable in reports and any future outputs when the findings of the study are disseminated *

◯ yes

◯ no

11. I agree to take part in this study *

◯ yes

◯ no

### Demographics

12. What is your role in the inpatient unit? *

- ◯ Nursing
- ◯ Health care assistant
- ◯ medical
- ◯ Patient and family support team
- ◯ Occupational therapy/physiotherapy team
- ◯ Management
- ◯ Administration
- ◯ Other
- ◯ Other

13. How long have you worked on the inpatient unit in Marie Curie Hospice Liverpool *

- ◯ Less than 6 months
- ◯ 6-12 months
- ◯ 12 months - 3 years
- ◯ 3 years -5 years
- ◯ 5 years - 10 years
- ◯ 10 years +

### Confidence

14. On a scale of 1-10 how confident would you feel to implement a FEWS tool as part of inpatient care? *

| 1 | 2 | 3 | 4 | 5 | 6 | 7 | 8 | 9 | 10 |
|---|---|---|---|---|---|---|---|---|---|

least confident most confident

15. How-on a scale of 1-10 how confident would you feel to review a FEWs tool as part of inpatient care? *

| 1 | 2 | 3 | 4 | 5 | 6 | 7 | 8 | 9 | 10 |
|---|---|---|---|---|---|---|---|---|---|

least confident most confident

### Barriers

16. What are the barriers to FEWS being used on the ward? (Select one or more option) *

- [ ] Confidence in  the FEWs tool working as a falls prevention tool
- [ ] Unware how to access the electronic version of the FEWS tool
- [ ] Not aware of what the FEWS tool is
- [ ] Lack of time
- [ ] Other
- [ ] Feel this isn't needed
- [ ] There are no barriers to using FEWs
- [ ] Lack of printed forms available
- [ ] Not aware of how to use the FEWS tool

17. How do you feel that the barriers  to implementing FEWs could be overcome? (Select one or more option) *

- [ ] Training on the use of FEWs
- [ ] Improved staffing levels
- [ ] Tool more accessible (admin to support printing out forms)
- [ ] Multidisciplinary involvement in completing the FEWS tool
- [ ] Written guidance on how to complete
- [ ] Other

18. If you answered 'other' to question 17, please provide details below

### Education

19. Have you received training on FEWS? *

- ◯ Yes
- ◯ No

20. How did you access training on FEWs?
(You can select more than one answer)

- ☐ Hospice education sessions
- ☐ 1:1 training during clinical practice
- ☐ MDT discussions
- ☐ Journal clubs
- ☐ Other

21. If you answered 'other' to the previous question, please provide details below

### Current practice

The FEWS tool is part of a multi-dimension model for falls management which aims to help the healthcare provider to determine the level of supervision a patient may require.

22. Who do you think should be responsible for initiating the FEWs tool? completing and reviewing the tool?
(You can select more than one answer) *

- ☐ Nursing staff
- ☐ Physiotherapist
- ☐ Other
- ☐ Medical staff
- ☐ Health care assistants
- ☐ Occupational therapist

23. If you answered 'other' to the previous question, please provide details below

24. Who do you think should be responsible for completing the FEWs tool?
(You can select more than one answer) *

- ☐ Occupational therapist
- ☐ Health care assistants
- ☐ Physiotherapist
- ☐ Medical staff
- ☐ Nursing staff
- ☐ Other

25. If you answered 'other' to the previous question, please provide details below

26. Who do you think should be responsible for reviewing the level of observation required using the FEWs tool?
(You can select more than one answer) *

- ☐ Occupational therapist
- ☐ Nursing staff
- ☐ Health care assistants
- ☐ Physiotherapist
- ☐ Other
- ☐ Medical staff

27. If you answered 'other' to the previous question, please provide details below

28. Do you know when FEWs should be initiated? *

- ○ Unsure
- ○ On admission
- ○ Following a fall or near miss
- ○ If medical condition detoriates
- ○ When risk of fall is identified

This content is neither created nor endorsed by Microsoft. The data you submit will be sent to the form owner.

Microsoft Forms
