## Supplementary file - Interview guide for "The implementation of a falls observational tool in a palliative care setting: a mixed methods study"

**FEWS Interview Guide**

*Thank you for taking the time to participate in an interview to discuss the Falls Early Warning Score (FEWs). I am going to start the interview by asking three demographic questions. I will then give you a little introduction to what FEWs is and the purpose of this study.*

What is your role?

How long have you been in this role?

Do you routinely complete FEWs as part of your role?

*FEWS is a tool which can be used to assess the level of observation a patient admitted to a hospice inpatient unit might need to reduce their risk of falls. This tool looks to guide nursing staff to identify an appropriate level of supervision for patients based on individual needs. FEWS aims to support plans of care for vulnerable individuals as set out in the Marie Curie patient falls policy. There are currently no validated tools to assess the level of observation required to reduce the risk of falls in palliative and end of life care patients. The purpose of this project is to explore and understand how an improvement of staff confidence and engagement in using a falls observational falls assessment tool could impact on the incidence of falls in a hospice inpatient setting.*

*The interview questions have been created around responses we received for a questionnaire about FEWs.*

*Firstly, I want to talk about how accessible the FEWs chart is in practice.*

- What is the most popular way to fill in FEWs on the IPU? (eg paper or electronic?)
    - Can you describe the advantages of this?
    - Can you describe the disadvantages of this?
    - How might this be made easier for you?

- What other ways might FEWs be completed? (eg: electronic)
    - 
    - Can you describe the advantages of this?
    - Can you describe the disadvantages of this?

**FEWS Interview Guide**

*I want to move on to talk about some of the challenges related to FEWs described in the questionnaire. The first is around time taken to complete FEWs.*

- Can you describe how much time it takes to complete the initial FEWs assessment?
  - What are the challenges to this?
  - What are the positives of completing the assessment?
  - Can you give any examples in practice?

- Can you describe how much time it takes to complete the regular FEWs assessments?
  - What are the challenges to this?
  - What are the positives of completing the regular assessments?
  - Can you give any examples in practice?

*One of the important areas identified in the questionnaire was around education. I just want to explore this a bit further with you.*

- Have you personally received any education around FEWs?
  - If yes, how did you find this?

- What type of education would be most useful for you to use FEWs in your daily practice? (eg: in person presentation/1:1 sessions/written resources)
  - How do you think this would look/work in clinical practice?
  - Why do you think this would be a useful way to help you use FEWs better?

- How would you feel about a 'step by step' guide to accompany FEWs
  - What should be in a step-by-step guide?
  - What would help visually?
  - Where would this be kept?

*I want to further explore completion of FEWs and whose responsibility this might be. It was highlighted in the questionnaire that it is important to consider how a whole MDT approach might be considered.*

- How do you think other members of the MDT should be involved in FEWs?
  - Who do you think this should be?
  - How do you think this should happen?
- Do you think that communication between different teams is important with FEWs?
  - Can you explain why?
  - How do you think this could be improved?

- How might FEWs be incorporated into MDT meetings?

- What do you think might be the benefits of this?
- What might be the drawbacks?

*The overall aim of FEWs is to improve patient care by reducing the incidence of falls in the hospice IPU setting. We want to explore this further.*

- Can you describe any examples, from clinical practice, of an incident of a patient falling?
    - How did this make you feel?
    - How did it impact the patient?
    - How do you feel the use of FEWs might have changed this scenario?

- How would you feel if FEWs was made an essential part of admission assessments?
    - How might this help you/your team?
    - What might this mean for patient care?
