## Supplementary file - FEWS Early Warning Score Form for "The implementation of a falls observational tool in a palliative care setting: a mixed methods study"

Name ..........................................
D.O.B..........................................
NHS/EMIS number..............................

### Falls Early Warning Score (FEWS)

#### PATIENT OBSERVATION CHART

Using the patient's information relating to their clinical presentation, staff should calculate a total score.
This score should then be used as a **guide** to assess the level of observation the patient requires.
Scores should be calculated at least daily and reviewed if patients medical, physical or cognitive condition changes or following a fall or assistance required to prevent a fall.

**It is Important to remember that this is a recommendation and nurses should use their clinical judgement when finally deciding on an appropriate level of observation.**

| CLINICAL PRESENTATION | SCORE |
|---|---|
| Confused/agitated | +1 |
| Unsteady when standing/mobilising | +1 |
| Falls history <6/52 | +1 |
| Urinary/faecal urgency | +1 |
| Postural hypotension | +1 |
| Inpatient fall during admission | +1* |
| **Medically unstable*** | +1 |
| **Environmental risk factors*** | +1 |
| **Fatigue/breathlessness*** | +1 |
| **Completely immobile/unconscious*** | -3 |
| **Compliant with safety/mobility advice & recommendation*** | -3 |
| **Other*** | +/- |

| LEVEL OF OBSERVATION | | SCORE |
|---|---|---|
| LEVEL A | Minimum 2 hourly | >0 |
| LEVEL B | Minimum hourly | 1-2 |
| LEVEL C | Minimum 30mins | 3-5 |
| LEVEL D | Constant 1:1 supervision | >6 |

- Adapted by Colette Parfitt & Emma Kirk October 2020 Marie Curie Hospice Liverpool from; Avoiding falls level of observation assessment tool (AFLOAT) Northumbria Healthcare NHS foundation trust Oct 2019

Name ..........................................
D.O.B..........................................
NHS/EMIS number..........................

| Date | Time | score | Level of Observation | Comments | Initials |
|---|---|---|---|---|---|
| | — | | | | |

- Adapted by Colette Parfitt & Emma Kirk October 2020 Marie Curie Hospice Liverpool from; Avoiding falls level of observation assessment tool (AFLOAT) Northumbria Healthcare NHS foundation trust Oct 2019

Name ............................................
D.O.B............................................
NHS/EMIS number..............................

| Date | Time | score | Level of Observation | Comments | Initials |
|---|---|---|---|---|---|
| | — | | | | |

- Adapted by Colette Parfitt & Emma Kirk October 2020 Marie Curie Hospice Liverpool from; Avoiding falls level of observation assessment tool (AFLOAT) Northumbria Healthcare NHS foundation trust Oct 2019

Name ........................................
D.O.B.........................................
NHS/EMIS number.............................

| Date | Time | score | Level of Observation | Comments | Initials |
|---|---|---|---|---|---|
| | — | | | | |

- Adapted by Colette Parfitt & Emma Kirk October 2020 Marie Curie Hospice Liverpool from; Avoiding falls level of observation assessment tool (AFLOAT) Northumbria Healthcare NHS foundation trust Oct 2019

Name ...........................................
D.O.B............................................
NHS/EMIS number...............................

- Adapted by Colette Parfitt & Emma Kirk October 2020 Marie Curie Hospice Liverpool from; Avoiding falls level of observation assessment tool (AFLOAT) Northumbria Healthcare NHS foundation trust Oct 2019
